# Comparison of Screening Guidelines for Cardiovascular disease Prevention and Early Detection: *a scoping review*

**DOI:** 10.1101/2023.08.05.23293697

**Authors:** Mohammed Abd ElFattah Mohammed Darwesh Badawy, Lin Naing, Nik Ani Afiqah Tuah

## Abstract

**Objective:** Globally, cardiovascular disease (CVD) has a significant role in morbidity and early death. This review’s objective is to provide a summary of the strengths and weaknesses in the variety of screening guidelines made by several international organizations for the early identification and prevention of CVD.

**Methods:** As the reporting guideline for this review, we used Preferred Reporting Items for Systematic Reviews and Meta-Analyses Extension for Scoping Reviews (PRISMA-ScR). We performed a scoping review using a few guideline-specific databases. We tabulated the main key differences between the included screening guidelines for CVD prevention and early detection from different perspectives.

**Results:** We included a total of 20 unique guidelines that were developed by various organizations throughout the world and focused on screening for CVD prevention and early detection out of the 2,466 guidelines discovered by our search based on our eligibility criteria. We concluded that the target populations, CVD risk assessment techniques, outcomes, and categories of the chosen CVD prevention guidelines widely varied. Additionally, some guidelines adopted no grading scheme for their evidence, while others did not advocate the use of any imaging screening tools in their evaluation of the CVD risk.

**Conclusions:** This scoping review highlights the areas of each guideline’s strengths and weaknesses and conducts a systematic comparison of a number of worldwide guidelines for CVD prevention and early diagnosis.

**What is already known in this review?:** Based on the most recent evidence and consensus among experts, each nation creates its own set of guidelines for the early detection and prevention of CVD.

**What this review adds:** This review conducted a systematic comparison and summarized the strengths and weaknesses of the various screening guidelines made by numerous international organizations for the early detection and prevention of CVD.

**• How this review might affect research, practice or policy:**

- This review provided opportunities to improve the future development of the clinical practice guidelines for CVD prevention and early detection.

## INTRODUCTION

The primary cause of disability and early mortality worldwide is cardiovascular disease (CVD). The prevalence of cardiovascular disease (CVD), which is expected to reach 130 million patients by 2035 with related mortality of about 24 million deaths, could result in healthcare sector costs of more than a trillion dollars worldwide [1,2]. Heart or blood vessel disorders such as coronary artery disease (CAD), cerebrovascular disease, peripheral arterial disease, and deep vein thrombosis are all considered to fall within the category of CVD [3]. The primary pathology in CVD is atherosclerosis, which develops and increases with age and typically manifests as acute coronary and cerebrovascular events that occur unexpectedly and frequently result in death before receiving the necessary medical care [4].

The term “cardiovascular disease (CVD) prevention” describes a systematic plan of measures aimed at minimizing and eliminating the disabilities associated with CVDs. These actions may be focused at the community level or the individual level [5]. All national organizations and joint societies in the healthcare sector are most concerned with ensuring that the recommended guideline for CVD preventive measures is properly and thoroughly implemented because the reduction of CVD prevalence and, consequently, CVD-related deaths, is the ultimate goal of CVD prevention [6,7].

Population-targeted prevention techniques emphasize altering lifestyle choices regardless of individual CVD risk to reduce the population’s total exposure to CVD risk factors. Contrarily, individual-based preventative techniques focus on high-risk populations to delay the beginning of CVD by reducing personal risk factors [8]. The term “CVD risk” refers to the possibility of experiencing fatal or nonfatal CVD events, such as a myocardial infarction or stroke in the foreseeable future [4]. Individual CVD risk prevention options include the “vertical” approach, which aims to manage a single risk according to predefined cut-offs regardless of the presence of concomitant risk factors or the “total” cardiovascular risk approach to preventing CVD is dependent on the individual’s odds of having fatal or nonfatal CVD events in a predetermined period concerning the presence of several related risk factors rather than a single risk factor [9].

Based on the most recent research and by evaluating the evidence-based, each nation or joint society creates a clinical practice guideline for managing CVD preventative screenings [5]. Numerous methods for controlling CVD risk can stop both fatal and non-fatal CVD events. Therefore, the determination of the risk of any such CVD event should serve as the basis for any guideline’s recommendations for a particular method for the prevention of CVD [4].

These guidelines are accompanied by several paper risk-prediction charts and online risk calculators that enable management to be targeted under straightforward risk calculations of the anticipated CVD event. Many risk calculators have been created to estimate an individual’s total risk of developing CVD or to specifically evaluate one of the major CVD event risks, such as the World Stroke Organization-endorsed Stroke Riskometer [10]. Every national healthcare system ought to implement a CVD risk calculator that is more practical, accurate, and user-friendly and is tailored to the populace based on many crucial features, such as variables, predictive accuracy, discrimination index, applicability, understandability, and cost-effectiveness [11].

Various health organizations and societies have developed recommendations in these guidelines for the management of major CVD risk factors and preventing CVD events risk through a series of lifestyle modification recommendations, a protocol for particular screening tests, and numerous prophylactic drug therapies tailored to each CVD risk category. These recommendations offer a framework that has been approved for the creation of national advice on the prevention of CVD risk in their community, taking into account the unique political, economic, social, and medical situations [4].

The goal of this scoping review is to draw attention to the differences between these guidelines from several perspectives, including the strength of the recommendations and the level of evidence substantiating them, risk assessment tools, the risk categories and risk outcomes associated with the target populations, as well as the advised non-invasive screening tests in each guideline.

## METHODS OF SCOPING REVIEW

### Study and search strategy

The National Guideline Clearinghouse (United States), the National Library for Health Guidelines Finder (United Kingdom), the Canadian Medical Association Clinical Practice Guidelines InfoBase and The GIN international guideline library were used in our scoping review to look for guidelines for the screening of CVDs. The search included the years 2000 through 2022, and only English-language guidelines were among the outcomes. The main search keywords are “cardiovascular disease,” “CVD prevention,” “cardiovascular disease prevention,” “cardiovascular disease screening,” “CVD screening,” “CVD screening guideline,” “cardiovascular risk screening,” “screening guideline,” and “guideline.”

### Study selection

We utilized Preferred Reporting Items for Systematic reviews and Meta-Analyses extension for Scoping Reviews (PRISMA-ScR) in this scoping review (**Figure 1**) [13], and completed the special reporting checklist for scoping reviews (**Appendix 1**). PRISMA-ScR checklist). In the initial literature search of this scoping review, 2,466 guidelines were found. All duplicates, partial guidelines, guidelines written by unidentified organizations, commentaries, guidelines not specifically focused on CVD, and other unrelated guidelines were excluded. Only 20 of the 40 full-text guidelines we assessed were eligible for inclusion in this review; the remaining guidelines were either not focused on screening CVD risks or were limited to a particular CVD disease.

**Figure 1.**
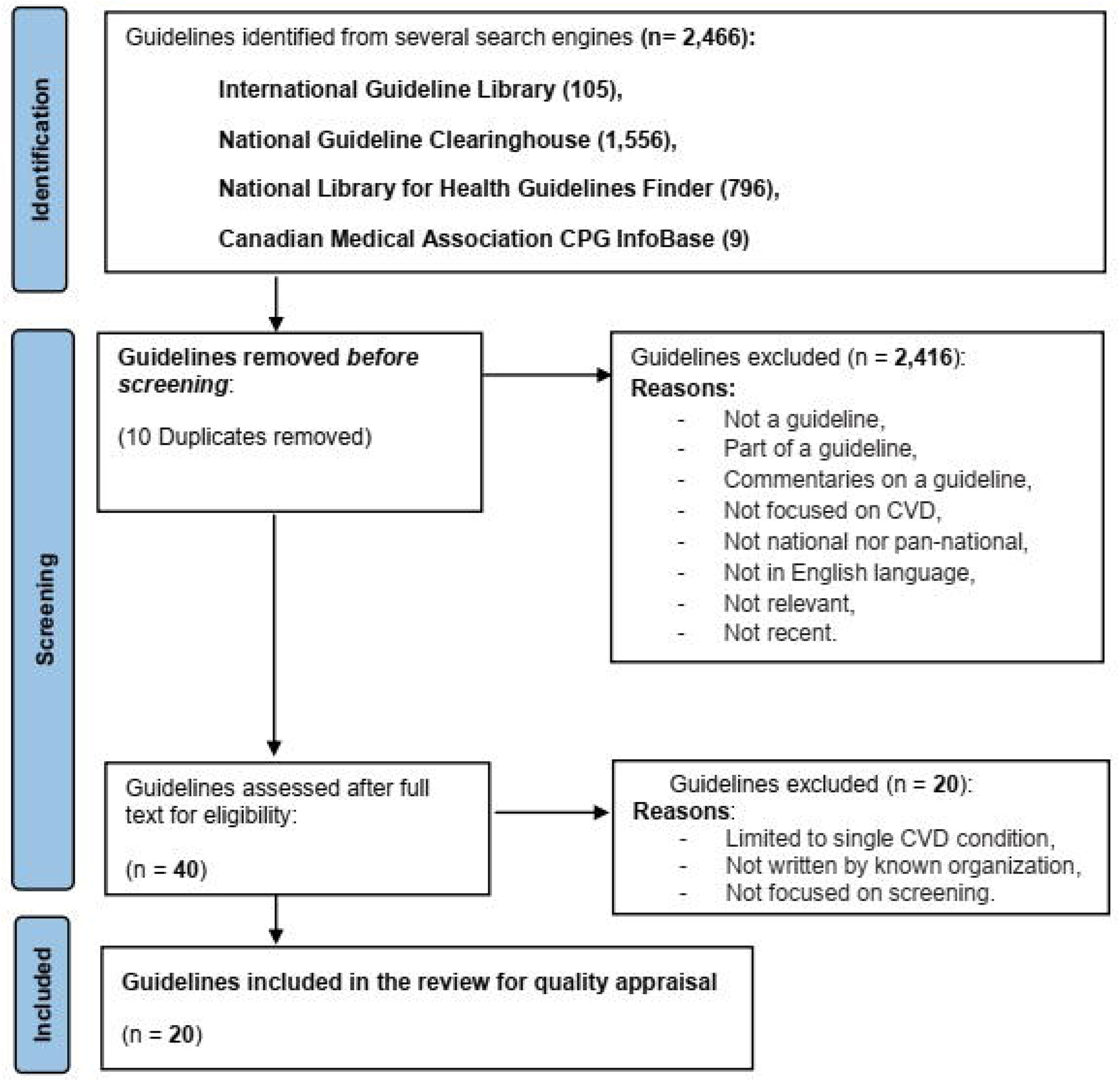
Identification of guidelines via databases

### Data extraction and tabular comparison

We also tabularly presented the main key differences between the included screening guidelines for CVD prevention and early detection in this review from different perspectives including the guideline’s recommendations grading system, to compare the “**level of evidence”** which represents an obvious strategy for conveying the quality of evidence to clinicians and “**the strength of the recommendation”**, which is defined as the extent to which one can be confident that the desirable consequences of an intervention outweigh its undesirable consequences [14]. Additionally, we discuss the differences in the included guidelines in terms of their definition of the target population, their advocated CVD risk assessment tool, the risk factors they screened and how they characterize their risk assessment outcome and the associated risk categories. Also, we tabulated the common non-invasive imaging screening tools utilized by different guidelines as well as the various screening strategies employed by those guidelines. Each of these areas of comparison highlights the points of the strengths and weaknesses of each included CVD prevention guideline.

## RESULTS

The list of 20 screening guidelines for CVD prevention and early detection in **Table 1** includes the guidelines’ titles, the year they were published, the organizations involved in their creation and evaluation, and the regions where they should be used most effectively. Eight of them were created in Europe (two continental, two in Scotland, and four in the UK), Five in North America (four in the United States of America and one in Canada), three in Asia (one each in Singapore, Malaysia, and Brunei Darussalam), two in Australia, one in New Zealand, and one was a global collaboration.

**Table 1.**
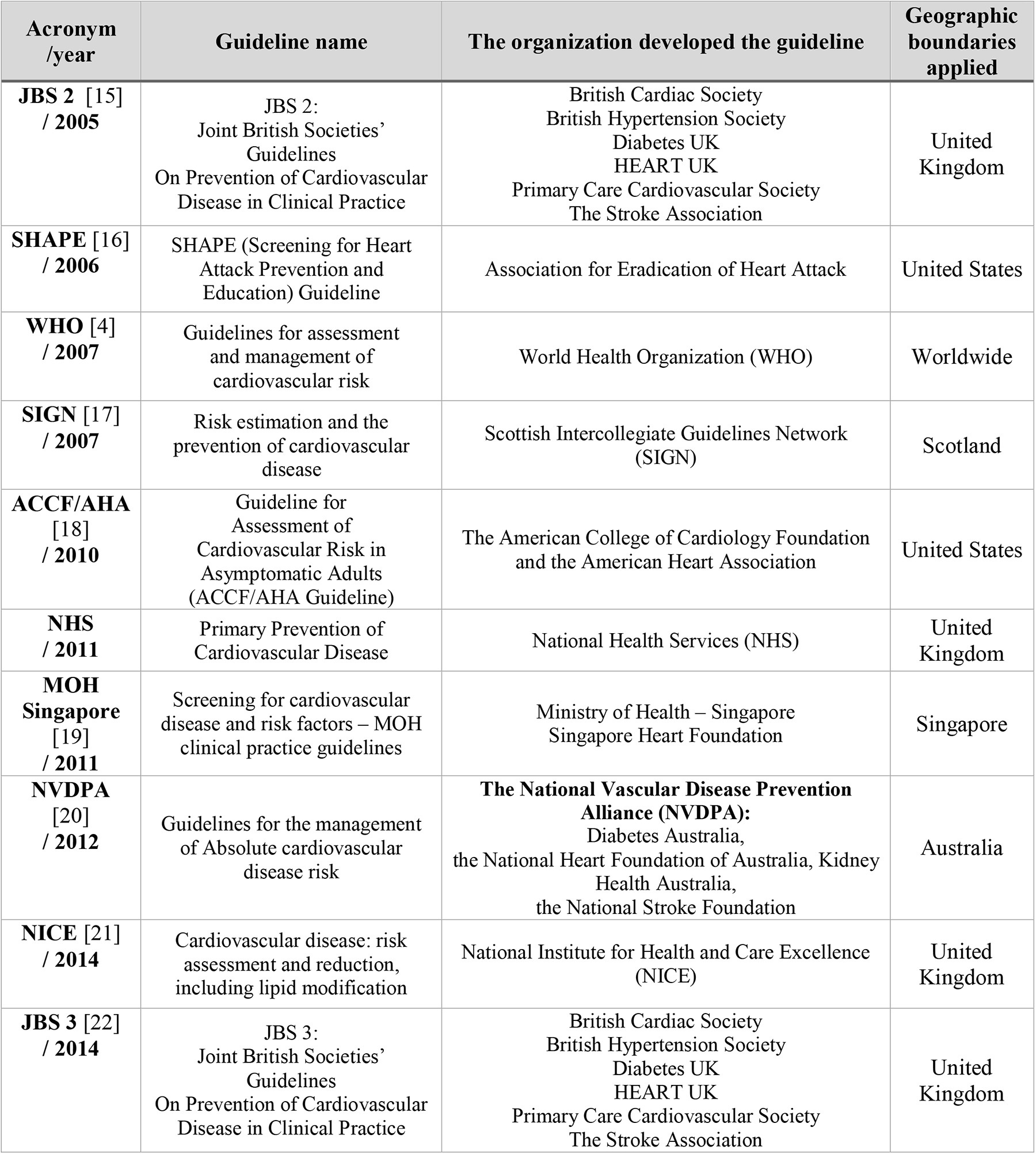
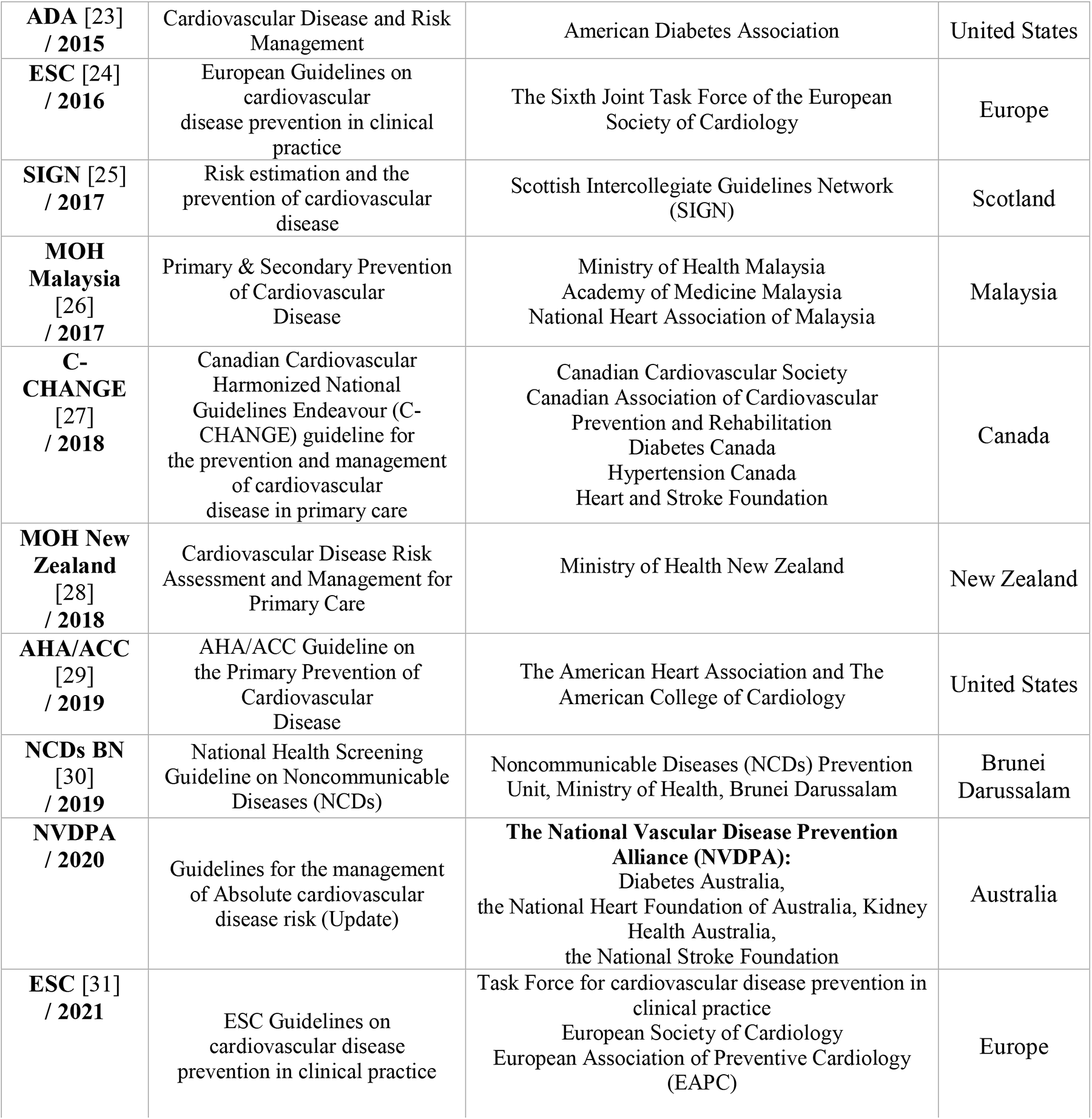
List of included screening guidelines for CVD prevention.

| **Acronym**<br>**/year** | **Guideline name** | **The organization developed the guideline** | **Geographic**<br>**boundaries**<br>**applied** |
| --- | --- | --- | --- |
| **JBS 2 [15]**<br>**/ 2005** | JBS 2:<br>Joint British Societies'<br>Guidelines<br>On Prevention of Cardiovascular<br>Disease in Clinical Practice | British Cardiac Society<br>British Hypertension Society<br>Diabetes UK<br>HEART UK<br>Primary Care Cardiovascular Society<br>The Stroke Association | United<br>Kingdom |
| **SHAPE [16]**<br>**/ 2006** | SHAPE (Screening for Heart<br>Attack Prevention and<br>Education) Guideline | Association for Eradication of Heart Attack | United States |
| **WHO [4]**<br>**/ 2007** | Guidelines for assessment<br>and management of<br>cardiovascular risk | World Health Organization (WHO) | Worldwide |
| **SIGN [17]**<br>**/ 2007** | Risk estimation and the<br>prevention of cardiovascular<br>disease | Scottish Intercollegiate Guidelines Network<br>(SIGN) | Scotland |
| **ACCF/AHA**<br>**[18]**<br>**/ 2010** | Guideline for<br>Assessment of<br>Cardiovascular Risk in<br>Asymptomatic Adults<br>(ACCF/AHA Guideline) | The American College of Cardiology Foundation<br>and the American Heart Association | United States |
| **NHS**<br>**/ 2011** | Primary Prevention of<br>Cardiovascular Disease | National Health Services (NHS) | United<br>Kingdom |
| **MOH**<br>**Singapore**<br>**[19]**<br>**/ 2011** | Screening for cardiovascular<br>disease and risk factors – MOH<br>clinical practice guidelines | Ministry of Health – Singapore<br>Singapore Heart Foundation | Singapore |
| **NVDPA**<br>**[20]**<br>**/ 2012** | Guidelines for the management<br>of Absolute cardiovascular<br>disease risk | **The National Vascular Disease Prevention**<br>**Alliance (NVDPA):**<br>Diabetes Australia,<br>the National Heart Foundation of Australia, Kidney<br>Health Australia,<br>the National Stroke Foundation | Australia |
| **NICE [21]**<br>**/ 2014** | Cardiovascular disease: risk<br>assessment and reduction,<br>including lipid modification | National Institute for Health and Care Excellence<br>(NICE) | United<br>Kingdom |
| **JBS 3 [22]**<br>**/ 2014** | JBS 3:<br>Joint British Societies'<br>Guidelines<br>On Prevention of Cardiovascular<br>Disease in Clinical Practice | British Cardiac Society<br>British Hypertension Society<br>Diabetes UK<br>HEART UK<br>Primary Care Cardiovascular Society<br>The Stroke Association | United<br>Kingdom |
| **ADA [23]**<br>**/ 2015** | Cardiovascular Disease and Risk Management | American Diabetes Association | United States |
| **ESC [24]**<br>**/ 2016** | European Guidelines on cardiovascular disease prevention in clinical practice | The Sixth Joint Task Force of the European Society of Cardiology | Europe |
| **SIGN [25]**<br>**/ 2017** | Risk estimation and the prevention of cardiovascular disease | Scottish Intercollegiate Guidelines Network (SIGN) | Scotland |
| **MOH Malaysia [26]**<br>**/ 2017** | Primary & Secondary Prevention of Cardiovascular Disease | Ministry of Health Malaysia<br>Academy of Medicine Malaysia<br>National Heart Association of Malaysia | Malaysia |
| **C-CHANGE [27]**<br>**/ 2018** | Canadian Cardiovascular Harmonized National Guidelines Endeavour (C-CHANGE) guideline for the prevention and management of cardiovascular disease in primary care | Canadian Cardiovascular Society<br>Canadian Association of Cardiovascular Prevention and Rehabilitation<br>Diabetes Canada<br>Hypertension Canada<br>Heart and Stroke Foundation | Canada |
| **MOH New Zealand [28]**<br>**/ 2018** | Cardiovascular Disease Risk Assessment and Management for Primary Care | Ministry of Health New Zealand | New Zealand |
| **AHA/ACC [29]**<br>**/ 2019** | AHA/ACC Guideline on the Primary Prevention of Cardiovascular Disease | The American Heart Association and The American College of Cardiology | United States |
| **NCDs BN [30]**<br>**/ 2019** | National Health Screening Guideline on Noncommunicable Diseases (NCDs) | Noncommunicable Diseases (NCDs) Prevention Unit, Ministry of Health, Brunei Darussalam | Brunei Darussalam |
| **NVDPA / 2020** | Guidelines for the management of Absolute cardiovascular disease risk (Update) | **The National Vascular Disease Prevention Alliance (NVDPA):**<br>Diabetes Australia,<br>the National Heart Foundation of Australia, Kidney Health Australia,<br>the National Stroke Foundation | Australia |
| **ESC [31]**<br>**/ 2021** | ESC Guidelines on cardiovascular disease prevention in clinical practice | Task Force for cardiovascular disease prevention in clinical practice<br>European Society of Cardiology<br>European Association of Preventive Cardiology (EAPC) | Europe |

The various grading systems that the included guidelines adopted are shown in **Table 2** to guarantee the quality of the evidence substantiating the recommendations and the strength of these recommendations. The Grading of Recommendations Assessment, Development and Evaluation (**GRADE**) is adopted by the Canadian guideline (C-CHANGE). The Scottish Intercollegiate Guidelines Network **(SIGN)** grading system was approved by the Ministry of Health Singapore, and Scottish guidelines. SIGN and GRADE grading systems were both used to rate the evidence and grade the recommendations of the WHO guideline.

**Table 2.** Guidelines’ Evidence and Recommendation Grading Systems.

| Guide / System | Level of the Evidence | Strength of the Recommendation |
| --- | --- | --- |
| WHO [4]<br>C-CHANGE [27] | **High ⊕⊕⊕⊕:** Further research is very unlikely to change our confidence in the estimate of | **Strong for ↑↑:** the panel considers that the desirable effects of adhering to a |
| / **GRADE system**<br>[32] | effect.<br>**Moderate** ⊕⊕⊕○: Further research is likely to have an important impact on our confidence in the estimate of effect and may change the estimate.<br>**Low** ⊕⊕○○: Further research is very likely to have an important impact on our confidence in the estimate of effect and is likely to change the estimate.<br>**Very low** ⊕○○○: We are very uncertain about the estimate. | recommendation clearly outweigh the undesirable effects<br>**Strong against** ↓↓: vice versa.<br>**Weak for** ↑: The panel considers that the desirable effects of adhering to a recommendation exceed the undesirable effects, although there is uncertainty.<br>**Weak against** ↓: vice versa. |
| ESC 2016 [24]<br>ESC 2021 [31]<br>MOH Malaysia [26]<br>/ **ESC system** | **A:** Data from multiple RCTs in the meta-analysis.<br>**B:** Data from a single RCT or from large non-randomized studies.<br>**C:** Consensus of opinion of experts and/or small studies, retrospective studies, and registries. | **Class I:** Evidence and/or general agreement that a certain diagnostic/treatment procedure is beneficial, useful and effective.<br>**Class II:** Conflicting evidence and/or divergence of opinion about the usefulness/effectiveness of the treatment.<br>-**Class IIa:** The weight of the evidence/opinion is in favour of utility/effectiveness.<br>-**Class IIb:** Utility/effectiveness is less established by evidence/opinion.<br>**Class III:** Evidence or general agreement that the treatment is not useful/effective and in some cases, may be harmful |
| AHA/ACC [29]<br>ADA [23]<br>/ **AHA/ACC system** | **A:** Evidence coming from RCTs and meta-analysis from several populations.<br>**B:** Evidence from a limited group of populations and a single RCT or non-randomized clinical trial.<br>**C:** Evidence from a very limited number of populations and consensus or expert opinions, reports and case series groups. | **Class I. Benefit Risk >>> (should be administered):** Evidence and/or general agreement that a treatment or procedure is beneficial, useful and effective.<br>- **Class IIa. Benefit >> Risk (REASONABLE to administer):** Contradictory evidence and/or divergent opinion on the benefit of the procedure, but the evidence supports that the treatment/procedure can help the patient.<br>- **Class IIb. Benefit ≥ Risk (MAY BE CONSIDERED):** Contradictory evidence and/or diverging opinion on the benefit of the procedure and is not well defined if the treatment/procedure can help the patient.<br>**Class III. Risk ≥ Benefit (SHOULD NOT be administered):** Evidence or general agreement that the treatment or procedure is given. It is not useful/effective, and in some cases, it can be harmful. |
| WHO [4]<br>SIGN 2007[17]<br>SIGN 2017[25]<br>MOH Singapore [19] | **1++:** High-quality meta-analysis, systematic reviews of RCTs or RCTs with very low risk of bias<br>**1+:** Well-conducted meta-analyses, systematic reviews or RCTs with low risk of bias<br>**1-:** Meta-analyses, systematic reviews or RCTs | **A:** At least one meta-analysis, systematic review or RCT rated as 1++ directly applicable to the target population; or a body of evidence consists mainly of 1+ studies, directly applicable to the target population and demonstrating the general consistency of the results. |
| /<br>**SIGN system** | with a high risk of bias<br>**2++:** High-quality systematic reviews of case-control or cohort studies<br>**2+:** Well-conducted cohort or case-control studies, with a low risk of confusion or bias and a moderate<br>**2-:** Case-control studies or cohorts with a high risk of confusion or bias and a significant risk that the relationship is not causal<br>**3:** non-analytical studies such as case reports or case series<br>**4:** Opinion of experts. | **B:** A body of evidence that includes 2++ studies, directly applicable to the target population and demonstrating overall consistency of results; or extrapolated evidence from 1++ or 1+ studies.<br>**C:** A body of evidence that includes 2+ studies, directly applicable to the target population and demonstrating overall consistency of results; or extrapolated evidence from 2++ studies.<br>**D:** Level of evidence 3 or 4; or extrapolated evidence from 2+ studies. |
| NICE [21]<br>NCDs BN [30]<br>/<br>**NICE system** | **I:** Evidence from meta-analyses, systematic reviews of randomized controlled trials (RCTs), or RCTs<br>**II:** Evidence from systematic reviews of case-control or cohort studies or any case control or cohort studies<br>**III:** Evidence from non-analytic studies, e.g., case reports, case series<br>**IV:** Expert opinion, formal consensus | None |
| NVDPA [20]<br>/<br>**National Health and Medical Research Council (NHMRC) system** [33] | **A:** Body of evidence can be trusted to guide practice<br>**B:** Body of evidence can be trusted to guide practice in most situations<br>**C:** The body of evidence provides some support for the recommendation but care should be taken in its application<br>**D:** The body of evidence is weak and recommendations must be applied with caution. | None |

The European Society of Cardiology (**ESC**) grading system has been approved by the European guidelines and the Malaysian Ministry of Health. American guidelines developed by the American College of Cardiology/American Heart Association (AHA/ACC) and American Diabetic Association (ADA) adopted **AHA/ACC** grading system.

The National Institute for Health and Care Excellence (**NICE**) levels of evidence served as the basis for the British guidelines and the National Health Screening Guideline that the Ministry of Health of Brunei Darussalam has adopted. The National Health and Medical Research Council (**NHMRC**) grading method was preferred by Australian guidelines [33].

However, none of the published guidelines produced by the Screening for Heart Attack Prevention and Education (SHAPE) task force, the Ministry of Health of New Zealand, or The Joint British Societies (JBS) explicitly outlined a grading system in their guidelines handbooks.

**Table 3** provides a thorough comparison between the included CVD screening guidelines in regard to defining the target population for each guideline, the CVD risk assessment tool used, CVD risk outcome and categories chosen.

**Table 3.** Comparison between the included CVD prevention guidelines regarding population, risk outcome, categories, factors and assessment tool.

| Acronym /Year | Target population | Risk outcome | Risk categories | Risk factors | Risk assessment tool |
| --- | --- | --- | --- | --- | --- |
| **JBS 2 [15] / 2005** | <u>All those people who are at **high risk**:</u><br>with any form of established atherosclerotic CVD.<br>without established CVD but who have a combination of risk factors_CVD risk of > 20% over 10 years)<br>with diabetes mellitus (type 1 or 2).<br><u>Other people with an **elevated single risk factor**:</u><br>elevated blood pressure > 160 mm Hg systolic or > 100 mm Hg diastolic.<br>total cholesterol to HDL ratio > 6.0<br>familial dyslipidemia<br>with a family history of premature CVD. | The total risk of developing CVD (coronary heart disease (CHD) and stroke) over 10 years | CVD risk < 10% over 10 years.<br>CVD risk 10 - 20% over 10 years.<br>CVD risk > 20% over 10 years | age, sex, smoking habit, systolic blood pressure, and the ratio of total cholesterol to HDL cholesterol. | **Joint British Societies' cardiovascular disease (CVD) risk prediction chart** |
| **SHAPE [16] / 2006** | Noninvasive screening of all asymptomatic men 45 to 75 years of age and asymptomatic women 55 to 75 years of age (except those defined as very low risk). | Risk of subclinical atherosclerosis | Very low risk.<br>Lower risk.<br>Moderate risk.<br>Moderately high risk.<br>High risk<br>Very High risk. | coronary artery calcium score (CACs), carotid artery intima-media thickness (CIMT), ankle-brachial index (ABI), C-reactive protein (CRP) | **Atherosclerosis test** |
| **WHO [4] / 2007** | Individuals with asymptomatic atherosclerosis, based on their estimated total CVD risk (high-risk group). | the 10-year risk of combined myocardial infarction and | Green <10%<br>Yellow 10% to <20%<br>Orange 20% to | age, sex, blood pressure, presence or absence of | **WHO/ISH CVD risk charts** |
|  | Patients who already have symptoms of atherosclerosis, such as angina or myocardial infarction, transient ischemic attack, or stroke are the top priority in clinical practice for prevention efforts. | stroke risk (fatal and non-fatal). | <30%<br>Red 30% to <40%<br>Deep Red > 40% | diabetes, smoking status, and cholesterol level. |  |
| **SIGN [17] / 2007** | <u>Individuals should have an assessment of CVD risk at least every five years:</u><br>all adults aged 40 years or above,<br>individuals at any age with a first-degree relative who has premature atherosclerotic CVD or familial dyslipidemia.<br><u>People should be assumed to be at **high risk** (a 10-year CVD risk $\geq 20\%$) who:</u><br>have had a previous cardiovascular event (angina, myocardial infarction, stroke, transient ischemic attack or peripheral arterial disease)<br>with diabetes (type 1 or 2) over the age of 40 years.<br>with familial hypercholesterolemia. | **ASSIGN score**<br>(The ten-year percentage risk of developing cardiovascular disease (any manifestation of coronary heart disease or cerebrovascular disease including transient ischemic attacks) in those who are disease-free at recruitment). | ASSIGN score below 20 is not currently high risk,<br>ASSIGN 20 or more is a high risk | age, sex, lifetime smoking habit (and number of cigarettes smoked per day), family history of CVD, socioeconomic status, blood pressure, weight and waist circumference, total cholesterol and HDL, glucose and renal function. | **The ASSIGN score online calculator** (ASsessing cardiovascular risk using SIGN guidelines to ASSIGN preventive treatment) |
| **ACCF/AHA [18] / 2010** | Initial assessment of the apparently healthy adult, beginning at age 20, for risk of developing CVD events associated with atherosclerotic vascular disease.<br>Excludes from consideration patients with a diagnosis of CVD, a coronary event, with known peripheral artery disease and cerebral vascular disease. | The 10-year risk of coronary heart disease (CHD) (MI and CHD death) | **Low** if the FRS is less than 10%,<br>**Moderate** if it is 10% to 19%,<br>**High** if it is 20% or higher | Age, sex, total cholesterol, HDL cholesterol, smoking, systolic blood pressure, antihypertensive medications | **Global risk scores** (e.g., the Framingham Risk Score) |
| **NHS / 2011** | <u>CVD 10-year risk assessments should be offered every 5 years to people with:</u><br>Hypertension.<br>a first-degree relative with premature cardiovascular disease (<60 years).<br>a first-degree relative with familial hypercholesterolemia (FH).<br>those who are selected for Keep Well health checks. | **ASSIGN score**<br>(The ten-year percentage risk of developing cardiovascular disease (any manifestation of coronary heart disease or cerebrovascular disease including transient ischemic attacks) in those who are disease-free at recruitment). | ASSIGN score below 20 is not currently high risk,<br>ASSIGN 20 or more is a high risk | age, sex, lifetime smoking habit (and number of cigarettes smoked per day), family history of CVD, socioeconomic status, blood pressure, weight and waist circumference, total cholesterol and HDL, glucose and renal function. | **The ASSIGN score online calculator** (ASsessing cardiovascular risk using SIGN guidelines to ASSIGN preventive treatment) |
| **MOH Singapore [19] / 2011** | Initial assessment of the apparently healthy adult, beginning at age 20, in Singapore | Estimate of person's 10-year CHD risk | **Low** if less than 10%,<br>**Moderate** if it is 10% to 19%,<br>**High** if it is 20% or higher | age, sex, total and HDL cholesterol levels, smoking status and systolic blood pressure | **Framingham-based NCEP ATP III** (National Cholesterol Education Program's Adult |
|  |  |  |  |  | Treatment Panel) modified according to Singapore CVD survey data. |
| **NVDPA [20] / 2012** | All adults aged 45–74 years who are not known to have CVD or to be at clinically determined high risk.<br>Aboriginal and Torres Strait Islander adults aged $\geq 35$ years | A probability of a CVD event within **5 years.** | **Low risk** (< 10%)<br>**Moderate risk** (10 to 15%)<br>**High risk** (> 15%) | age, gender, systolic blood pressure, smoking status, total and HDL cholesterol, Diabetes, ECG LVH | **Australian Absolute CVD Risk Calculator** |
| **NICE [21] / 2014** | adults who are at risk of or who have cardiovascular disease (CVD), such as heart disease and stroke up to and including age 84 years | 10-year QRISK®2 score + QRISK® Heart Age | 10-year risk of CVD is less than 10%<br>10-year risk of CVD is 10% or more. | Age, Gender, Ethnicity, Relevant family history, Smoking status, Systolic blood pressure, HDL, Total Cholesterol, BMI, Deprivation score, Diabetes, hypertensive medication, Rheumatoid arthritis, chronic kidney disease, Atrial Fibrillation | **QRISK2** risk assessment tool |
| **JBS 3 [22] / 2014** | JBS3 risk calculator to estimate both 10-year risk and lifetime risk of CVD in all individuals except for those with existing CVD or certain high-risk diseases: that is, diabetes age >40 years, patients with chronic kidney disease (CKD) stages 3–5, or familial hypercholesterolemia (FH). | the total risk of developing CVD on **short-term (10 years)** and **over the lifetime** + **Heart Age** | CVD risk < 10% over 10 years.<br>CVD risk 10 - 20% over 10 years.<br>CVD risk > 20% over 10 years | age, sex, ethnic group, BMI, smoking habit, systolic blood pressure, total cholesterol, HDL, diabetes, blood pressure treatment, close relative under 60 suffer from CVD, chronic kidney disease, atrial fibrillation, rheumatoid arthritis | **JBS3 risk online calculator** |
| **ESC [24] / 2016** | Adults >40 years of age, unless they are automatically categorised as being at high-risk or very high-risk based on documented CVD, DM (>40 years of age), kidney disease or highly elevated single risk factor. | The 10-year risk of only fatal CVD in the population at high CVD risk. | **Low risk:** < 1 %.<br>**Moderate risk:** 1 % 2 % 3 – 4 %.<br>**High risk:** 5 – 9 %<br>**Very High**<br>**risk:**<br>**10 – 14 %**<br>**15 % and**<br>**over** | age, sex, smoking, systolic blood pressure, total cholesterol | **SCORE** (Systematic Coronary Risk Estimation) risk charts |
| **SIGN [25]**<br>**/ 2017** | <u>Individuals with the following risk factors should be considered at high risk of CVD events:</u><br>established cardiovascular disease,<br>stage 3 or higher chronic kidney disease or micro- or macroalbuminuria,<br>familial hypercholesterolemia,<br>who are over the age of 40 and have diabetes,<br>who are under the age of 40 and have diabetes, and<br>at least 20 years duration of disease, or<br>target organ damage (e.g., proteinuria, micro- or macroalbuminuria, proliferative retinopathy or autonomic neuropathy), or<br>significantly elevated cardiovascular risk factors. | **ASSIGN score**<br>(The ten-year percentage risk of developing cardiovascular disease (any manifestation of coronary heart disease or cerebrovascular disease including transient ischemic attacks) in those who are disease-free at recruitment). | ASSIGN score below 20 is not currently high risk,<br>ASSIGN 20 or more is a high risk | age, sex, smoking, systolic blood pressure, total cholesterol, HDL cholesterol, family history of premature CVD, diagnosis of diabetes, diagnosis of rheumatoid arthritis and deprivation | **The ASSIGN score online calculator** (ASsessing cardiovascular risk using SIGN guidelines to ASSIGN preventive treatment) |
| **MOH Malaysia [26]**<br>**/ 2017** | These guidelines are developed to prevent CVD (heart disease and strokes) in all individuals in Malaysia. | **<u>Very High Risk</u>**<br>A FRS-CVD score that confers a 10-year risk for CVD of >30%<br>Established CVD<br>Diabetes mellitus with proteinuria<br>CKD with glomerular filtration rate (GFR) <30 µl/ min-1/ 1.73 m2 (Stage ≥4)<br>**<u>High Risk</u>**<br>Have a FRS-CVD score that confers a 10-year risk for CVD of >20%<br>Diabetes mellitus without target organ damage<br>CKD with GFR >30 - <60 µl/ min-1/ 1.73 m2 (Stage 3)<br>Very high levels of individual risk factors (LDL-C >4.9 mmol/L, BP >180/110 mmHg)<br>**<u>Intermediate (Moderate) Risk:</u>**<br>Have a FRS-CVD score that confers a 10-year risk for CVD of 10-20%<br>**<u>Low Risk:</u>**<br>Have a FRS-CVD score that confers a 10-year risk for CVD <10%. |  | **<u>Non-modifiable</u>** – increasing age, gender, family history of premature CVD, ethnicity.<br>**<u>Modifiable</u>** – diet and dietary patterns, smoking, physical inactivity, obesity/overweight, hypertension, dyslipidemia and pre-diabetes/diabetes. | **Framingham Risk Score** |
| **C-CHANGE [27]**<br>**/ 2018** | Canadian adults who have or are at risk of developing chronic cardiovascular diseases, including hypertension, diabetes, dyslipidemia, heart failure and stroke, and the risk factors for these conditions, including smoking, obesity and physical inactivity. | Estimate of person's 10-year CHD risk | **Low** if less than 10%,<br>**Moderate** if it is 10% to 19%,<br>**High** if it is 20% or higher | age, sex, total and HDL cholesterol levels, smoking status and systolic blood pressure | **Modified Framingham Risk Score** |
| **MOH New Zealand** | The target population includes all men without prior CVD aged 45– | Percentage of risk of a CVD | **< 3 %.**<br>**3 – 9 %.** | Age, gender, ethnicity, smoking, diabetes, systolic | **PREDICT CVDRA** |
| [28] / 2018 | 74 years, and all women without prior CVD aged 55–74 years.<br>It also includes Māori, Pacific or South-Asian peoples from an age 15 years younger than the starting age for the general population. | event within 5 years. | 10 – 14 %.<br>15+ % | blood pressure, treatment of BP, lipid-lowering treatment, antithrombotic treatment, total cholesterol, HDL, Family history of CVD, chronic kidney disease, atrial fibrillation, Deprivation quartile. | equations |
| AHA/ACC [29] / 2019 | Adults who are 40 to 75 years of age and are being evaluated for cardiovascular disease prevention should undergo a 10-year atherosclerotic cardiovascular disease (ASCVD) risk estimation. | 10-year ASCVD risk for patients aged 40-79 or Lifetime risk for patients aged 20-59. | **Low-risk** (<5%)<br>**Borderline risk** (5% to 7.4%)<br>**Intermediate risk** (7.5% to 19.9%)<br>**High risk** (≥20%) | Age, sex, race, systolic and diastolic blood pressure, total cholesterol, HDL, LDL, history of diabetes, smoking status, hypertension treatment, statin or aspirin therapy | **ASCVD Risk Estimator** |
| NCDs BN [30] / 2019 | Asymptomatic adult population 40 years and above<br>Consider screening below 40 years old if the presence of risk factor(s) | the 10-year risk of combined myocardial infarction and stroke risk (fatal and non-fatal). | **Low** < 10 %<br>**Intermediate** (10 to 20 %)<br>**High/Very High** > 20 % | age, sex, blood pressure, presence or absence of diabetes, smoking status, and cholesterol level. | **WHO/ISH CVD risk prediction chart for WPR-A** |
| ESC [31] / 2021 | **SCORE2:** apparently healthy people aged 40-69 years with risk factors that are untreated or have been stable for several years.<br>**SCORE2-OP:** competing risks in apparently healthy people aged ≥70 years. | **SCORE2:** 10-year risk of fatal and non-fatal CVD events (myocardial infarction, stroke).<br>**SCORE2-OP:** 5-year and 10-year fatal and nonfatal CVD events (myocardial infarction, stroke). | **Low-to-moderate CVD risk:**<br><50 years: <2.5%<br>50-69 years: < 5%<br>≥70 years: <7.5%<br>**High CVD risk:**<br><50 years: 2.5%- <7.5%<br>50-69 years: 5%- < 10%<br>≥70 years: 7.5% - < 15%<br>**Very high CVD risk:**<br><50 years: ≥7.5%<br>50-69 years: ≥ 10%<br>≥70 years: ≥15% | age, sex, smoking, systolic blood pressure, total cholesterol | **SCORE2** (Updated SCORE) & **SCORE2-OP** Updated SCORE for old people) |

**Table 4** lists the numerous non-invasive screening techniques and instruments that are advised by various CVD preventive guidelines. These screening tools include resting Electrocardiography (ECG), Echocardiography, Coronary Artery Calcium (CAC) obtained by Computed Tomography (CT), Carotid artery intima-media thickness (cIMT) assessment using ultrasonography, carotid artery ultrasound, abdominal aorta ultrasound, Ankle-brachial index, Treadmill ECG stress test, Stress myocardial perfusion imaging (MPI), Arterial stiffness commonly measured using either aortic pulse wave velocity (PWV) or arterial augmentation index, Coronary CT angiography or MRI.

Each guideline recommends a specific group of screening tools for specific CVD risk categories, which vary between different guidelines as indicated in the table. The low-risk category is indicated by the colour green, the intermediate-risk category by the colour yellow, and the high-risk category by the colour red.

**Table 4.**
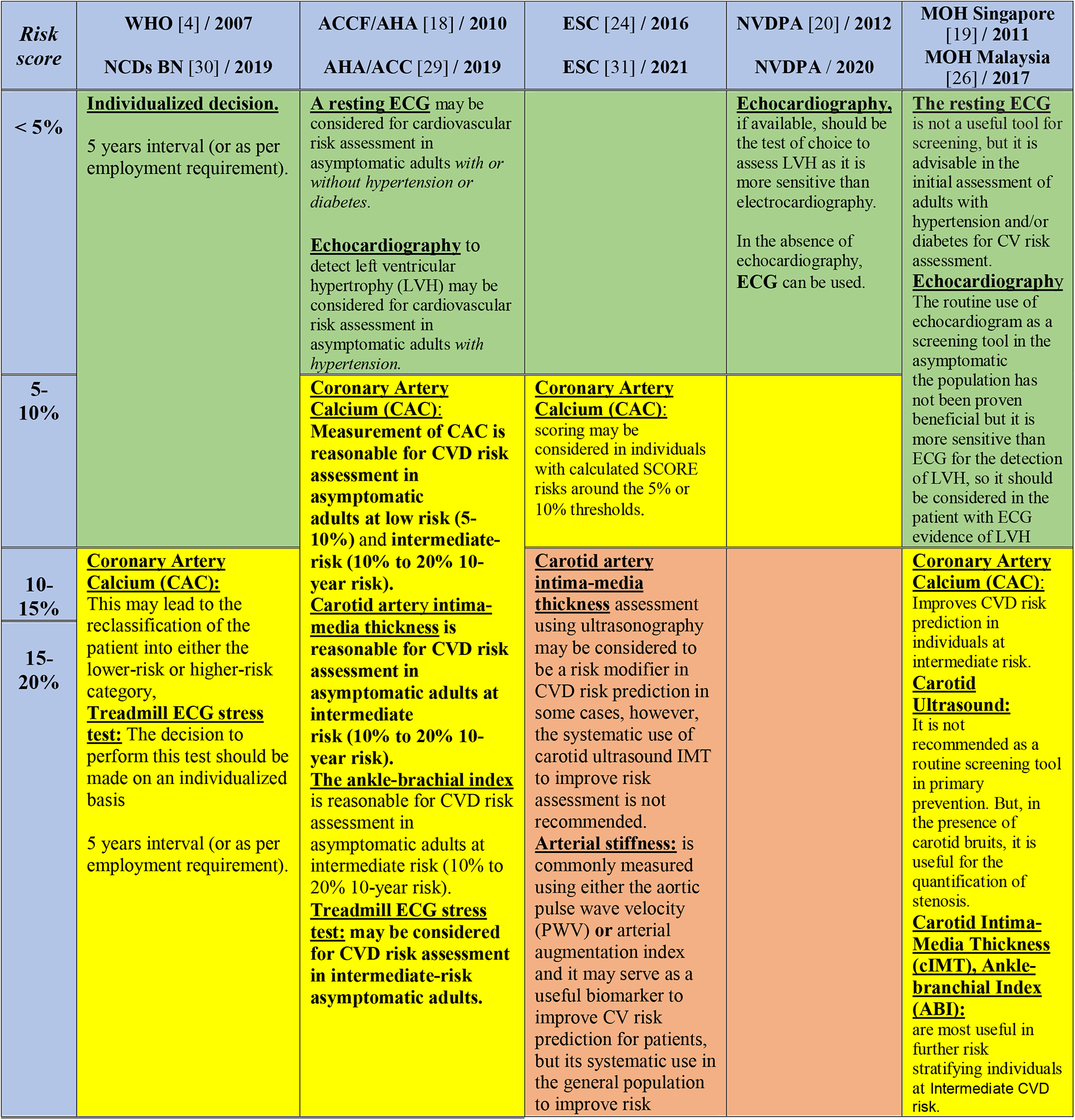
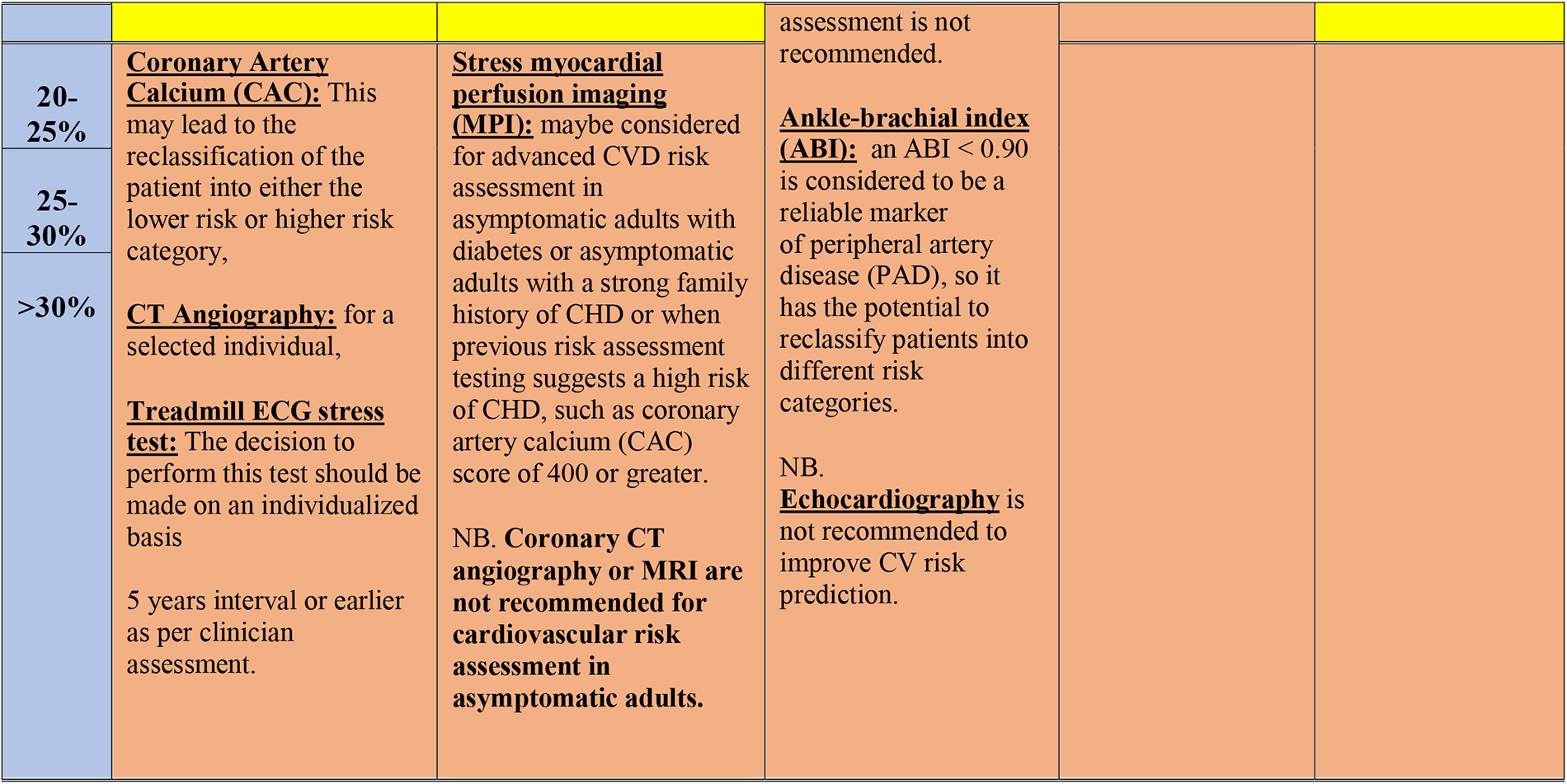
Differences in recommended imaging tools in the included CVD prevention guidelines.

## DISCUSSION

### Different Guidelines’ Evidence and Recommendation Grading Systems

Our results revealed in **Table 2** the diversity of the evidence and recommendation grading systems for the included CVD prevention guidelines as well as the deficiency of some guidelines from reporting clear system of grading their quality level of evidence and strength of their recommendations. These results are in concordance with literature as guideline developers around the world are inconsistent in how they rate the quality of evidence, describe clear criteria for the selection of evidence and grade the strength of recommendations, also; They frequently neglect to adequately consider the overall picture provided by a body of evidence as well as the methodological quality of each individual studies. As a result, it might be difficult for guideline users to comprehend the messages that grading systems are trying to convey [32,34].

Expert clinicians and organizations that provide recommendations to the clinical community frequently make errors because they do not consider sufficient account of the quality of evidence [35]. The adoption of grading systems by patients, physicians, and policymakers is facilitated by their simplicity in making assessments of the level of the evidence and the strength of the recommendations [36]. For those employing guidelines and recommendations, judgments will be more transparent if the criteria for grading the strength of the recommendations and evaluating the quality of the evidence are explicit and detailed [32]. In addition, The balance between health benefits, side effects, quality of evidence, applicability, and the certainty of the baseline risk should all be considered in judgments about the strength of recommendations and formulating the guidelines [37].

### Guidelines comparison regarding population, risk outcome, categories, factors and assessment tool

After data extraction and analysis, the selected CVD prevention guidelines vary in defining their target populations appointed for efforts of screening and early detection of CVD risk. Some guidelines target all individuals in the nation in an effort to the ultimate prevention of CVD risk such as Malaysian and Singaporean guidelines. Others define a specific age range for screening of CVD risk, while European guideline (ESC 2021), for instance, updated a specific CVD risk screening tool, SCORE2-OP, for elderly people. The majority of the included guidelines recommend the initial CVD risk assessment for apparently healthy, asymptomatic adults. However, some simply took into account patients with a high CVD risk to mitigate resources utilization in healthcare settings with limited resources, such as Scottish guidelines created by SIGN. Additionally, patients with a diagnosis of established CVD, a coronary event, known peripheral artery disease, or cerebral vascular disease are excluded by the American guidelines developed by the AHA/ACC.

Although assessing CVD risk is essential for identifying the necessity for preventative treatment as well as defining the intensity of treatment [38–40], evidence indicates that healthcare professionals frequently underestimate a patient’s CVD risk [41]. The authors of the guidelines recommend using a variety of risk assessment tools to assist healthcare professionals in estimating the risk of the first cardiovascular event in adult patients, [42] including risk prediction charts like the WHO/ISH risk prediction chart and the JBS2 risk prediction charts as well as online calculators like the Framingham Risk Score (FRS), QRISK^®2^ (version two of the QRISK^®^ CVD risk algorithm), Assessing Cardiovascular Risk using Scottish Intercollegiate Guidelines Network (ASSIGN), Systematic Coronary Risk Evaluation (SCORE), JBS3 risk online calculator and Australian Absolute CVD_JRisk Calculator.

Each of these risk assessment tools categorizes the population into distinct CVD risk categories defining the risk outcome as the ten-year percentage risk of developing cardiovascular diseases such as any manifestation of coronary heart disease or cerebrovascular disease including transient ischemic attacks. However, some guidelines, such as those from Australia and New Zealand, would rather advise estimating five-year risk (as opposed to ten-year risk), as both risk and risk management can change significantly over ten years, and as a result, predicting a five-year risk is more likely to be useful in actual practice. They also argued that the majority of randomized controlled trials of CVD preventive medications are based on five years or fewer of treatment, hence the best estimates of treatment benefits are over five rather than ten years [20,28].

On the other hand, certain guidelines, such as those created by JBS3 and NICE, emphasize the lifetime risk of CVD events to include a sizable population of persons who have a low 10-year risk of a CVD event but a high lifetime event risk [21,22].

### Different recommended screening tools for CVD prevention

A variety of non-invasive screening methods have been researched and are being used more frequently in clinical settings to detect CVD, including the resting electrocardiogram (ECG), which has been used since the late 1800s to diagnose CVD and is frequently used to evaluate the risk of CVD in asymptomatic adults with or without diabetes or hypertension [43]. Additionally, Echocardiography, which is typically used to detect left ventricular hypertrophy (LVH) in asymptomatic adults with hypertension, is more sensitive than an ECG in detecting LVH. However, some guidelines suggest that routine use of echocardiography as a screening tool in the asymptomatic population has not been proven to be beneficial [19,26].

Another example is the Coronary Artery Calcium Score (CAC) obtained by computed tomography (CT), which enhances CVD risk prediction in those at intermediate CVD risk and may result in the patient being reclassified into either the lower risk or higher risk category. Additionally, measuring the carotid intima-media thickness (cIMT) using ultrasound may be thought of by some guidelines as a risk modifier in the prediction of CVD risk in specific circumstances, but its routine application to enhance risk assessment is not usually advised. Also, routine screening methods for primary prevention such as abdominal aorta ultrasound and carotid artery ultrasound are typically not advised. However, they are helpful for quantifying stenosis when bruits are present.

It should be emphasized that some guidelines only adopted targets for the high-risk patients’ lifestyle, blood pressure, lipids, and glucose levels in addition to cardiovascular protective medication therapy for particular clinical indications. These guidelines did not recommend any imaging screening tool in their assessment of the CVD risk. These guidelines include “Joint British Societies’ Guidelines on Prevention of Cardiovascular Disease in Clinical Practice” (JBS2), “Risk Estimation and The Prevention of Cardiovascular Disease” (SIGN 2007, 2017), “Cardiovascular Disease Risk Assessment and Management for Primary Care” by Ministry of Health New Zealand and “Canadian (C-CHANGE) guideline for the prevention and management of cardiovascular disease in primary care”. Although JBS3 2014 improved its risk assessment model by incorporating the use of non-invasive imaging technologies to identify subclinical atherosclerosis, it noted that their use is not advised for CVD risk assessment in the primary preventive setting [22].

## CONCLUSION AND RECOMMENDATIONS

In conclusion, we highlighted several areas of differences, strengths, and weaknesses of many screening guidelines for CVD prevention and early detection.

To assure the Rigour of the development of the guidelines, techniques like the Grading of Recommendations Assessment, Development, and Evaluation (GRADE) methodology, which is one of the soundest approaches for evaluating the quality of a body of evidence in systematic reviews and clinical practice guidelines, provides a transparent and organized methodology for developing and presenting evidence summaries, rating the quality of the evidence and judging the strength of the recommendations [12,32]. Additionally, those making the recommendations must consider how the data will be implemented in a specific context, considering important factors that could modify the scope of the predicted effects [37]. In addition, it is recommended that policymakers should incorporate digital transformation strategies for CVD risk screening and the adoption of cutting-edge digital technology into their future guidelines for CVD early detection and prevention.

## STRENGTHS AND LIMITATIONS

Our scoping review has several advantages, including the capacity to develop a systematic comparison between different guidelines produced globally for CVD risk screening, prevention, and early detection and contrast the points of strengths and weaknesses of their recommendations. A thorough database search and reading of a sizable number of guidelines for CVD prevention and early detection were also conducted. A specified methodology and specific inclusion and exclusion criteria were used in our search.

However, there are a few potential limitations to this scoping review that should be considered. First, the rapid review methods and minimal bias assessment. Second, despite using a systematic approach of comparison and highlighting several strengths and weaknesses, this review’s comparison of CVD screening guidelines is not exhaustive, and many comparison points may be missing.

## Data Availability

All data produced in the present work are contained in the manuscript.

## DECLARATIONS

### Ethics approval and consent to participate

As this review is based only on published studies, ethics approval and consent to participate are not applicable.

### Consent for publication

Not applicable.

### Availability of data and materials

All data analyzed during this study and supporting its findings are included in this published article and all studies included in this review are available in **Table 1**.

### Authors’ contributions

All authors contributed toward databases search, drafting and critically revising the paper and agree to be accountable for all aspects of the work. The authors read and approved the final manuscript.

## Acknowledgements

This research is made possible through the generous support of the PAPRSB Institute of Health Sciences, Universiti Brunei Darussalam, Brunei Darussalam.

## ABBREVIATIONS

ABI: Ankle-branchial Index
ACCF/AHA: The American College of Cardiology Foundation and the American Heart Association
ADA: American Diabetes Association
AGREE II: Appraisal of Guidelines for Research and Evaluation II
AHA/ACC: The American Heart Association and The American College of Cardiology
ASSIGN: ASsessing cardiovascular risk using SIGN guidelines to ASSIGN preventive treatment
CAC: Coronary Artery Calcium
CAD: coronary artery disease
C-CHANGE: Canadian Cardiovascular Harmonized National Guidelines Endeavour
cIMT: Carotid Intima-Media Thickness
CPG: Clinical practice guideline
CT: Computed Tomography
CVD: Cardiovascular disease
CVDRA: CVD risk assessment
EAPC: European Association of Preventive Cardiology
ECG: electrocardiogram
ESC: European Society of Cardiology
GRADE: The Grading of Recommendations Assessment, Development and Evaluation
ISH: International Society of Hypertension
JBS: Joint British Societies’ Guidelines
MOH: Ministry of Health
MPI: myocardial perfusion imaging
NCDs: Noncommunicable Diseases
NCEP ATP: National Cholesterol Education Program’s Adult Treatment Panel
NHMRC: National Health and Medical Research Council
NHS: National Health Services
NICE: National Institute for Health and Care Excellence
NVDPA: The National Vascular Disease Prevention Alliance
PRISMA-ScR: Systematic reviews and Meta-Analyses extension for Scoping Reviews
PWV: aortic pulse wave velocity
SCORE: Systematic Coronary Risk Estimation
SCORE2-OP: Updated SCORE for old people
SHAPE: Screening for Heart Attack Prevention and Education
SIGN: Scottish Intercollegiate Guidelines Network
UK: United Kingdom
WHO: World Health Organization
WPR-A: Western Pacific Region A

